# Treatment Outcomes and Associated Factors Among Children with Epilepsy Attending the Pediatric Follow-Up Clinic at Hiwot Fana Comprehensive Specialized Hospital

**DOI:** 10.1101/2025.06.15.25329654

**Authors:** Mohammed Saleye, Hanan Abdirahman, Tesfaye Assabe, Temesgen Libe

**Affiliations:** Jigjiga University College of Medicine and Health Science, Jigjiga, Ethiopia; Haramaya university College of Health and Medical Science, Harar, Ethiopia

**Author notes:** **Corresponding Author:** Temesgen Libe, College of Health and Medical Sciences, Haramaya University, Harar, Ethiopia.

**Keywords:** Epilepsy, Epilepsy treatment outcome, Seizure control, Children, Ethiopia

## Abstract

**Background:** Epilepsy is a common neurological disorder that significantly affects children’s development, behavior, and quality of life. In low-resource settings, treatment outcomes remain suboptimal.

**Objectives:** This study assessed treatment outcomes and associated factors among children with epilepsy attending the pediatric follow-up clinic at Hiwot Fana Comprehensive Specialized Hospital in Eastern Ethiopia.

**Methodology:** A hospital-based cross-sectional study was conducted from November 5, 2020, to February 5, 2021, involving 140 children aged 6 months to 18 years who had been on antiepileptic drugs (AEDs) for at least six months. Data was collected through structured caregiver interviews and medical record reviews. Seizure control was categorized as good (seizure-free for at least six months) or poor (persistent seizures despite appropriate AED use). Adherence was measured using the Morisky 8-Item Medication Adherence Scale. Multivariable logistic regression was used to identify factors associated with treatment outcomes.

**Result:** Among the 140 participants, 59.3 percent had poor seizure control. Generalized seizures were the most common type (65.7 percent), and 78.6 percent were on monotherapy, with phenytoin being the most prescribed AED. Poor adherence to AEDs was observed in 46.4 percent of patients. In the adjusted analysis, seizure frequency greater than one per week before AED initiation (AOR = 11.036; 95% CI: 3.616–33.677; p < 0.001) and poor adherence to AEDs (AOR = 4.917; 95% CI: 2.452–9.861; p < 0.001) were significantly associated with poor treatment outcomes.

**Conclusion:** Nearly six in ten children had poor seizure control. High pre-treatment seizure frequency and poor adherence to medication were the main factors associated with poor outcomes. Interventions to promote early diagnosis and improve adherence through caregiver education and health system support are critical to improving epilepsy management in children.

## Introduction

Epilepsy is a common neurological disorder characterized by recurrent, unprovoked seizures. It can significantly impact cognitive development, behavior, and quality of life. A diagnosis of epilepsy is made when a person experiences two or more seizures that are not caused by temporary conditions such as fever, metabolic disturbances, or brain infections [1].

Epilepsy is a common neurological disorder characterized by recurrent, unprovoked seizures. It can significantly impact cognitive development, behavior, and quality of life. A diagnosis of epilepsy is made when a person experiences two or more seizures that are not caused by temporary conditions such as fever, metabolic disturbances, or brain infections [2].

The main goal of epilepsy treatment is to achieve seizure control. A good treatment outcome refers to complete or near-complete seizure control, while a poor outcome is defined by persistent or worsening seizures despite treatment. Studies suggest that about two-thirds of epilepsy cases can be well-controlled with antiepileptic drugs (AEDs) [3]. In low-resource settings like Ethiopia, phenobarbital is commonly used due to its low cost and wide availability. However, treatment failure still occurs in nearly one-third of patients [4, 5]

Although AEDs do not cure epilepsy, they can help most patients maintain long-term seizure control. Poor treatment outcomes can result from inappropriate drug selection, suboptimal dosing, adverse effects, poor adherence, or drug-resistant epilepsy. Understanding these factors is essential for optimizing patient care [6].

Several studies in LMICs, including Ethiopia, have shown suboptimal seizure control rates, with only 60–70% of patients achieving good outcomes [7, 8]. This has significant implications for children, as uncontrolled seizures can affect their development, education, and psychosocial well-being.

Despite the growing burden of pediatric epilepsy in Ethiopia, there is a lack of research on treatment outcomes and associated factors—particularly in the eastern part of the country.

This study aims to assess the treatment outcomes and associated factors among children with epilepsy attending the pediatric follow-up clinic at Hiwot Fana Specialized University Hospital in Harar, Eastern Ethiopia. To the best of our knowledge, this is the first study of its kind in this setting.

## Methods and Materials

A hospital-based cross-sectional study was conducted prospectively from November 5, 2020, to February 5, 2021, among children diagnosed with epilepsy and attending the pediatric follow-up outpatient department (OPD) at Hiwot Fana Comprehensive Specialized Hospital (HFCSH), Harar, eastern Ethiopia. HFCSH is a tertiary-level teaching hospital affiliated with Haramaya University and serves approximately 5.8 million people from the Harari Region, Dire Dawa City Administration, Somali Region, and Eastern Hararghe Zone of Oromia Regional State. Pediatric neurology follow-up clinics are conducted every Tuesday and Wednesday, where most patients are children with epilepsy, according to Health Management Information System (HMIS) data from 2019 (unpublished).

All children aged 6 months to 18 years with a clinical diagnosis of epilepsy, who had been on antiepileptic drugs (AEDs) for at least 6 months and were attending follow-up at the pediatric clinic during the study period, were considered eligible. Children younger than 6 months or those who had been on AEDs for less than 6 months were excluded.

A total of 140 children who met the inclusion criteria were consecutively enrolled using a non-probability convenience sampling method.

Data was collected through face-to-face interviews using a semi-structured questionnaire developed based on relevant literature. The questionnaire was initially prepared in English and later translated into Amharic and Afan Oromo. It was pre-tested on 5% of the expected sample size at Dilchora Hospital, and necessary adjustments were made. Additional clinical and treatment-related data were abstracted from the patients’ medical records concurrently during the study period, from November 5, 2020, to February 5, 2021.

The data collection process was carried out over a three-month period by two trained BSc nurses, under the supervision of one pediatric resident.

Treatment outcomes were classified based on seizure control status. Good seizure control was defined as being seizure-free for six months or more. Poor seizure control referred to the occurrence of one or more seizures per month over a six-month period despite adequate adherence and trials of at least two appropriately dosed AEDs either as monotherapy or in combination. Partial seizure control was defined as a reduction in seizure frequency by more than 50%, while complete seizure control referred to total remission of seizures for a minimum of six months [9].

Adherence to AEDs was measured using the Morisky 8-Item Medication Adherence Scale. A score of 0 indicated high adherence, scores of 1–2 indicated medium adherence, and scores greater than 2 were considered low adherence. Adherence was assessed based on the agreement between the patient’s or caregiver’s medication-taking behavior and the clinician’s prescription[10].

Descriptive statistics were used to summarize the socio-demographic, clinical, and treatment-related characteristics of the study participants. Bivariate logistic regression analysis was conducted to identify factors associated with treatment outcomes. Variables with a p-value of less than 0.20 in the bivariate analysis were subsequently entered into a multivariable logistic regression model to adjust for potential confounders. The strength of association was expressed using adjusted odds ratios (AORs) with corresponding 95% confidence intervals (CIs). Statistical significance was declared at a p-value less than 0.05. All data analyses were performed using Stata version 17.0.

## Ethical Consideration

Ethical approval was obtained from the Institutional Health Research Ethics Review Committee (IHRERC) of the College of Health and Medical Sciences, Haramaya University, prior to data collection with ethical approval number of IHRERC/241/2020. The study was conducted in full compliance with the ethical guidelines set forth by the IHRERC. Written, informed, and voluntary consent was obtained from the hospital administration, and an official letter of cooperation was submitted to Hiwot Fana Comprehensive Specialized Hospital (HFCSH). The outpatient department director was formally notified, and the hospital reserved the right to discontinue the study in the event of any misconduct or ethical violations.

Data extracted from medical records was anonymized to ensure confidentiality. Participants were informed of their right to decline or withdraw from the study at any stage without any repercussions.

Voluntary, informed, written, and signed consent was also obtained from the parents or caregivers of all participating children. Confidentiality was strictly maintained by excluding names and personal identifiers from the data collection tools.

This research adhered to the ethical principles outlined in the Declaration of Helsinki.

## Result

### Socio demographic characteristics of patients with epilepsy

The study included 140 patients diagnosed with epilepsy. More than half of the participants (69.3%) were male, and the majority (70%) were between 1 and 10 years of age. The most common age at the onset of epilepsy was between 1 and 5 years, accounting for 72 cases (52.4%). Additionally, 75 patients (53.6%) had been receiving antiepileptic drugs (AEDs) for less than two years. [Table 1]

**Table 1:** Socio demographic characteristics of children with epilepsy at HFCSH.

| Variable |  | Frequency (Percentage) |
| --- | --- | --- |
| Sex of the child | Male | 97 (69.3) |
|  | Female | 43 (30.7) |
| Age of the child | 6-12months | 4 (2.9) |
|  | 1-5years | 55 (39.3) |
|  | 5-10years | 43 (30.7) |
|  | 10-15years | 32 (22.9) |
|  | 15-18 years | 6 (4.3) |
| Residence | Urban | 68 (48.6) |
|  | Rural | 72 (51.4) |
| Primary care giver | Mother | 102 (72.9) |
| Father | Father | 20 (14.3) |
| Brother | Brother | 7 (5) |
| Sister | Sister | 7 (5) |
| Other | Other | 4 (2.9) |
| Marital status of the primary care giver | Single | 7 (5) |
|  | Married | 105 (75) |
|  | Divorced | 16 (11.4) |
|  | Widowed | 12 (8.6) |
| Educational status of primary care giver | Less than high school | 69 (49.3) |
|  | High school | 50 (35.7) |
|  | College and above | 21 (15.0) |
| Family in come | >1500 birr | 64 (45.7) |
|  | <=1500 | 76 (54.3) |

Regarding the socio-demographic characteristics of primary caregivers, mothers were reported as the primary caregivers in 102 cases (72.9%), while fathers assumed this role in only 20 cases (14.3%). Most caregivers were married (75%), and half (50%) had an educational level below high school. Furthermore, more than half of the caregivers (54.3%) reported a monthly household income of less than 1,500 birr. [Table 1]

### Clinical Profile, Antiepileptic Therapy, and Treatment Outcomes Among Pediatric Epilepsy Patients at HFCSH Follow-Up Clinic

Of the 140 pediatric epilepsy patients included in the study, 92 (65.7%) had generalized epilepsy, 42 (30%) had focal epilepsy, and 6 (4.3%) had unknown onset. A known cause of epilepsy was identified in 13 patients (9.3%), with pyogenic meningitis and stroke each accounting for 5 cases (3.6%). Comorbid conditions were reported in 15 patients (10.7%), while epilepsy syndromes were diagnosed in 10 patients (7.1%). Prior to the initiation of antiepileptic drug (AED) therapy, more than half of the participants (60.7%) experienced more than one seizure per week. [Table 2]

**Table 2:**
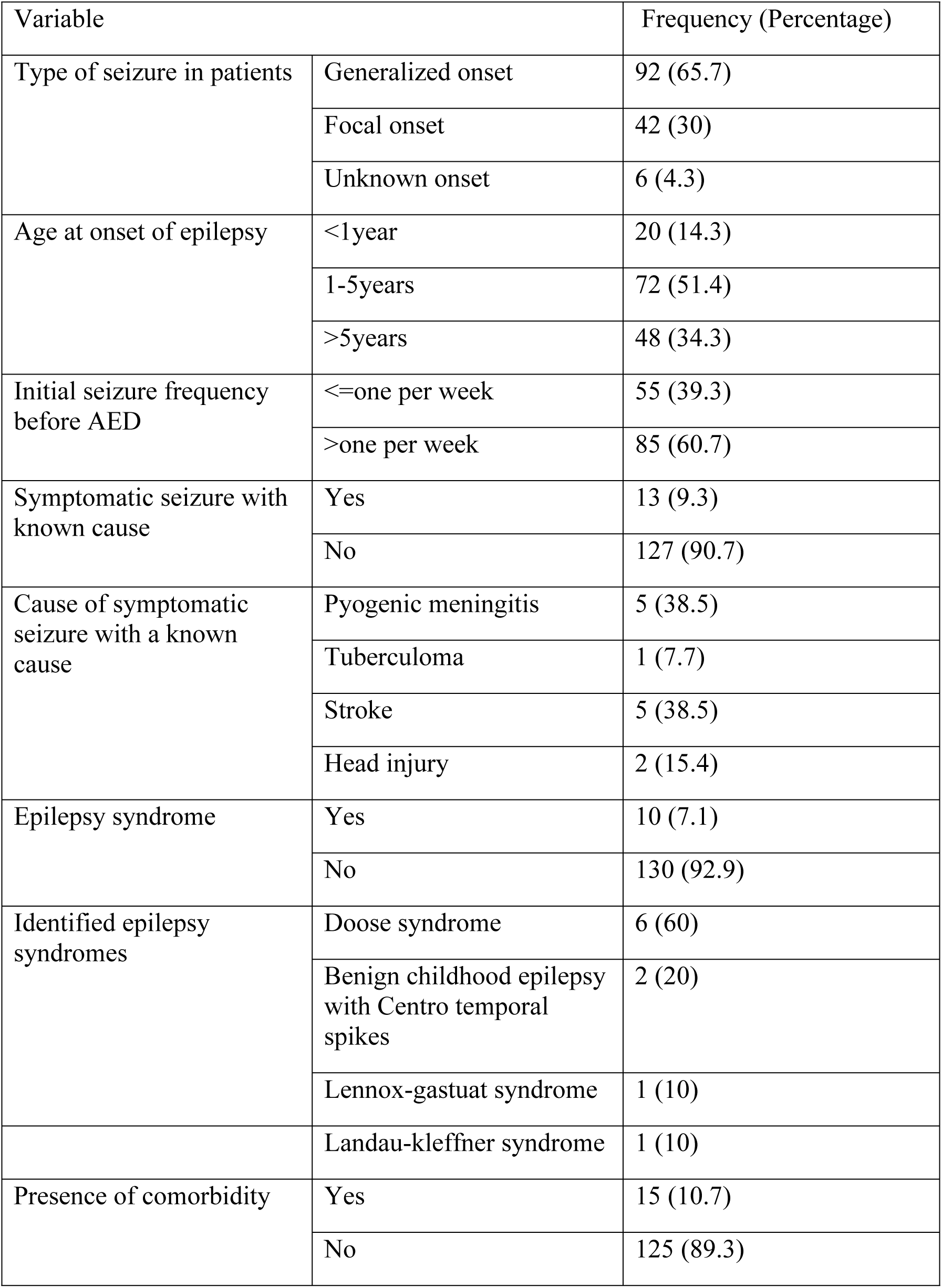
Clinical Characteristics of Pediatric Epilepsy Patients at HFCSH.

Most patients (110; 78.6%) were receiving monotherapy, with phenytoin being the most prescribed AED. Poor adherence to medication was noted in 65 patients (46.4%), and only one patient (0.7%) reported experiencing adverse effects related to AED use. In terms of seizure control, 57 patients (40.7%) achieved complete seizure control, 67 (47.9%) had partial control, and 16 (11.4%) had poorly controlled seizures. [Table 3]

**Table 3:** Antiepileptic Therapy and Treatment Outcomes Among Children with Epilepsy at HFCSH.

| Variable |  | Frequency (Percentage) |
| --- | --- | --- |
| Mode of therapy | Monotherapy | 110 (78.6) |
|  | Dual therapy | 26 (18.6) |
|  | Poly therapy | 4 (2.9) |
| Type of monotherapy prescribed | Phenytoin | 67 (47.9) |
|  | Phenobarbitone | 15 (10.7) |
|  | Valproate | 17 (12.1) |
|  | Carbamazepine | 11 (7.9) |
| Level of adherence | High level of adherence | 49 (35) |
|  | Medium level of adherence | 26 (18.6) |
|  | Low level of adherence | 65 (46.4) |
| Duration of epilepsy since initiation of AED | <2years | 75 (53.6) |
|  | 2-4years | 42 (30) |
|  | >4years | 23 (16.4) |
| Presence of adverse effect on AED | Gum hyperplasia | 1 (0.7) |
|  | No | 139 (139.3) |
| Seizure control status | Complete seizure control | 57 (40.7) |
|  | Partial seizure control | 67 (47.9) |
|  | Poor seizure control | 16 (11.4) |
| Outcome | Good outcome | 57 (40.7) |
|  | Poor outcome | 83 (59.3) |

### Factors associated with epilepsy treatment outcome among epileptic children at HFCSH pediatric follow up clinic

Treatment outcomes were categorized as good or poor based on clinical response to antiepileptic drug (AED) therapy.

Bivariate logistic regression analysis identified several variables with a p-value less than 0.20, making them eligible for inclusion in the multivariable model. These included residence, educational status of the primary caregiver, family monthly income, type of seizure, mode of AED therapy, level of adherence, seizure frequency before AED initiation, symptomatic seizures with a known cause, presence of comorbidities, and developmental delay. [Table 4]

**Table 4:** Bivariate analysis of factors associated with epilepsy treatment outcome at HFCSH pediatric follow up clinic.

| Variable | OR (95%CI) | P.value |
| --- | --- | --- |
| Residence | 2.13 (1.072 - 4.232) | **0.031** |
| Educational status of the primary care giver | 0.654 (0.049 – 1.045) | **0.076** |
| Family monthly in come | 2.962 (1.473 – 5.957) | **0.002** |
| Type of seizure | 0.602 (0.311 – 1.163) | **0.131** |
| Age at the onset of seizure | 1.027 (0.620 – 1.701) | 0.918 |
| Seizure frequency before AED | 22.150 (9.132 – 53.744) | **0.000** |
| Symptomatic seizure with known cause | 0.406 (0.106 – 1.545) | **0.186** |
| Comorbidity | 0.329(0.88-1.223) | **0.097** |
| Developmental delay | 0.266(0.56-1.261) | **0.095** |
| Microcephaly | 0.279 (0.317 – 2.450) | 0.249 |
| Mode of AED therapy | 1.440 (0.699 – 2.9720) | 0.323 |
| Level of adherence | 0.2(0.06-0.61) | **0.00** |

In the multivariable logistic regression analysis, only two variables were found to be significantly associated with epilepsy treatment outcomes. Children who experienced more than one seizure per week prior to initiating AED therapy had significantly higher odds of poor treatment outcomes compared to those with less frequent seizures (AOR = 11.036; 95% CI: 3.616–33.677; p < 0.001). Additionally, children with poor adherence to AED therapy were nearly five times more likely to have poor outcomes compared to those with good adherence (AOR = 4.917; 95% CI: 2.452–9.861; p < 0.001). Other variables, including residence, caregiver educational status, family income, seizure type, symptomatic etiology, comorbidities, and developmental delay, did not show statistically significant associations with treatment outcome in the adjusted analysis. [Table 5]

**Table 5:** Multivariate analysis of factors associated with epilepsy treatment outcome at HFCSH pediatric follow up clinic.

| Variable | AOR (95%CI) | P Value |
| --- | --- | --- |
| Residence | 1.724(0.308-9.543) | 0.53 |
| Educational status of the primary care giver | 0.489 (0.195 – 1.228) | 0.128 |
| Family monthly in come | 1.216 (0.310 – 4.766) | 0.779 |
| Type of seizure | 1.547 (0.501 – 4.775) | 0.448 |
| Seizure frequency before AED | 11.036 (3.616 – 33.677) | **0.000** |
| Symptomatic seizure with known cause | 0.617 (0.070 – 5.432) | 0.663 |
| Comorbidity | 0.298 (0.012 – 7.266) | 0.457 |
| Developmental delay | 1.155 (0.029 – 45.330) | 0.939 |
| Level of adherence | 4.917 (2.).452 – 9.861)) | **0.000** |

## Discussion

This study aimed to assess treatment outcomes and associated factors among children with epilepsy attending the pediatric follow up clinic at Hiwot Fana Comprehensive Specialized Hospital. We found that 59.3 percent of pediatric epilepsy patients had uncontrolled seizures. This finding contrasts with a study conducted in Gondar, Ethiopia, where 82 percent of children had controlled seizures [11]. The discrepancy may be due to differences in the definition of seizure control, as our study used a six-month seizure free period while the Gondar study used three months. However, our findings are consistent with reports from Ayder Comprehensive Specialized Hospital and Tikur Anbessa Specialized Hospital, where more than half of children had uncontrolled seizures [12, 13].

Two variables were independently associated with treatment outcomes: seizure frequency before initiation of antiepileptic drug therapy and adherence to prescribed medications.

Children who experienced more than one seizure per week before starting treatment were significantly more likely to have poor seizure control compared to those with less frequent seizures. The adjusted odds ratio was 11.036 with a confidence interval from 3.616 to 33.677 and a p value less than 0.001. This result is consistent with studies from Bangladesh and Finland which also found that high seizure frequency at baseline was a strong predictor of poor treatment outcomes [9, 14]. These findings highlight the importance of early diagnosis and prompt initiation of appropriate therapy to prevent the development of refractory epilepsy.

Adherence to antiepileptic drugs also played a critical role in determining treatment success. Children with poor adherence were nearly five times more likely to experience uncontrolled seizures compared to those with good adherence. The adjusted odds ratio was 4.917 with a confidence interval from 2.452 to 9.861 and a p value less than 0.001. This finding agrees with several studies conducted in Ethiopia and Nigeria, which identified nonadherence as a major challenge in the effective management of pediatric epilepsy [11, 12, 15]. Factors contributing to poor adherence may include caregiver misunderstanding of the treatment plan, limited access to medications, side effects, or missed clinic appointments. Interventions such as education, counseling, and medication reminders may improve adherence and seizure control.

Unlike findings from some previous studies, this study did not find significant associations between treatment outcomes and factors such as caregiver education level, residence, seizure type, symptomatic causes of epilepsy, or the presence of comorbidities [16, 17]. While lower caregiver education levels have been linked to adherence challenges in other settings, their lack of statistical significance in this study suggests that other healthcare and socio-economic factors may be influencing treatment success. Future research could further explore these dimensions to provide a more comprehensive understanding.

## Limitations

This study has several limitations. The cross-sectional design limits the ability to establish causal relationships between variables. The sample size was relatively small, which may reduce the generalizability of the findings. The absence of diagnostic tools such as electroencephalography and advanced neuroimaging limited accurate classification of seizure types and etiologies. In addition, data collection occurred during the COVID 19 pandemic, which may have affected follow up attendance and medication adherence. Despite these limitations, the study provides important insights into pediatric epilepsy management in a resource constrained setting.

## Conclusion

This study demonstrated that a significant proportion of children with epilepsy at Hiwot Fana Comprehensive Specialized Hospital had poor seizure control. Seizure frequency prior to treatment initiation and adherence to antiepileptic medication were the two major factors influencing treatment outcomes. These findings emphasize the importance of early identification of high-risk patients and implementation of adherence support strategies. Strengthening caregiver education, ensuring consistent drug availability, and improving clinic follow up systems are essential to enhancing epilepsy outcomes in children. Future studies should consider larger samples and prospective designs to provide stronger evidence for policy and clinical interventions.

## Data Availability

The data used to support the findings of this study are available from the corresponding author (Temesgen Teferi Libe) upon reasonable request to ensure patient confidentiality

## Data Sharing Statement

Any of the data used for analysis in the study is available from the corresponding author and ready to be provided up on reasonable request.

## Contributors

Manuscript conceptualization: M S, H A, T A, and T L; Data management and analysis: M S, H A, T A, and T L; investigation: M S, H A, T A, and T L; methodology: M S, H A, T A, and T L; Validation: M S, H A, T A, and T L; writing the original manuscript draft: M S and T L. All authors reviewed and provided inputs to the draft and approved the decision to submit for publication.

